# Influence of serosorting and intervention-mediated changes in serosorting on the population-level HIV transmission impact of pre-exposure prophylaxis among men who have sex with men: a mathematical modelling study

**DOI:** 10.1101/2020.02.26.20025700

**Authors:** Linwei Wang, Nasheed Moqueet, Anna Simkin, Jesse Knight, Huiting Ma, Nathan J. Lachowsky, Heather L. Armstrong, Darrell H. S. Tan, Ann N. Burchell, Trevor A. Hart, David M. Moore, Barry D. Adam, Derek R. MacFadden, Stefan Baral, Sharmistha Mishra

## Abstract

**Background:** HIV pre-exposure prophylaxis (PrEP) may change serosorting patterns. We examined the influence of serosorting on the population-level HIV transmission impact of PrEP, and how impact could change if PrEP users stopped serosorting.

**Methods:** We developed a compartmental HIV transmission model parameterized with bio-behavioural and HIV surveillance data among men who have sex with men in Canada. We separately fit the model with serosorting and without serosorting (random partner-selection proportional to availability by HIV-status (sero-proportionate)), and reproduced stable HIV epidemics (2013-2018) with HIV-prevalence 10.3%-24.8%, undiagnosed fraction 4.9%-15.8%, and treatment coverage 82.5%-88.4%. We simulated PrEP-intervention reaching stable coverage by year-1 and compared absolute difference in relative HIV-incidence reduction 10-year post-intervention (PrEP-impact) between: models with serosorting vs. sero-proportionate mixing; and scenarios in which PrEP users stopped vs. continued serosorting. We examined sensitivity of results to PrEP-effectiveness (44%-99%) and coverage (10%-50%).

**Findings:** Models with serosorting predicted a larger PrEP-impact compared with models with sero-proportionate mixing under all PrEP-effectiveness and coverage assumptions (median (inter-quartile-range): 8.1%(5.5%-11.6%)). PrEP users” stopping serosorting reduced PrEP-impact compared with when PrEP users continued serosorting: reductions in PrEP-impact were minimal (2.1%(1.4%-3.4%)) under high PrEP-effectiveness (86%-99%); however, could be considerable (10.9%(8.2%-14.1%)) under low PrEP effectiveness (44%) and high coverage (30%-50%).

**Interpretation:** Models assuming sero-proportionate mixing may underestimate population-level HIV-incidence reductions due to PrEP. PrEP-mediated changes in serosorting could lead to programmatically-important reductions in PrEP-impact under low PrEP-effectiveness (e.g. poor adherence/retention). Our findings suggest the need to monitor sexual mixing patterns to inform PrEP implementation and evaluation.

**Funding:** Canadian Institutes of Health Research

**RESEARCH IN CONTEXT:** *Evidence before this study:* We searched PubMed for full-text journal articles published between Jan 1, 2010, and Dec 31, 2017, using the MeSH terms “pre-exposure prophylaxis (PrEP)” and “homosexuality, male” and using key words (“pre-exposure prophylaxis” or “preexposure prophylaxis” or “PrEP”) and (“men who have sex with men” or “MSM”) in titles and abstracts. Search results (520 records) were reviewed to identify publications which examined the population-level HIV transmission impact or population-level cost-effectiveness of PrEP in high-income settings. We identified a total of 18 modelling studies of PrEP impact among men who have sex with men (MSM) and four studies were based on the same model with minor variations (thus only the most recent one was included). Among the 15 unique models of PrEP impact, three included serosorting. A total of nine models have assessed the individual-level behaviour change among those on PrEP and its influence on the transmission impact of PrEP. Specifically, the models examined increases in number of partners and reductions in condom use. Most models predicted that realistic increases in partner number or decreases in condom use would not fully offset, but could weaken, PrEP”s impact on reducing HIV transmission. We did not identify any study that examined the influence of serosorting patterns on the estimated transmission impact of PrEP at the population-level, or what could happen to HIV incidence if the use of PrEP changes serosorting patterns.

*Added value of this study:* We used a mathematical model of HIV transmission to estimate the influence of serosorting and PrEP-mediated changes in serosorting on the transmission impact of PrEP at the population-level among MSM. We found the impact of PrEP was higher under epidemics with serosorting, compared with comparable epidemics simulated assuming sero-proportionate mixing. Under epidemics with serosorting, when PrEP users stopped serosorting (while other men continue to serosort among themselves) we found a reduced PrEP impact compared with scenarios when PrEP users continued to serosort. The magnitude of reduction in PrEP impact was minimal if PrEP-effectiveness was high; however, could be programmatically-meaningful in the context of low PrEP-effectiveness (e.g., poor adherence or retention) and high PrEP coverage. To our knowledge, our study is the first to directly examine the influence of serosorting and PrEP-mediated changes in serosorting on the transmission impact of PrEP and its underlying mechanism.

*Implications of all the available evidence:* Our findings suggest that models which do not consider baseline patterns of serosorting among MSM could potentially underestimate PrEP impact. In addition to monitoring individual-level behavioural change such as condom use, our findings highlight the need to monitor population-level sexual mixing patterns and their changes over time among MSM in the design and evaluation of PrEP implementation.

## INTRODUCTION

Sexual mixing patterns - also known as patterns of “who has sex with whom” - influences the population-level transmission dynamics of sexually transmitted infections such as HIV.^1^ Mixing plays a role in how HIV may spread and persist, and thus how prevention interventions may fare at a population-level.^1^ However, the influence of mixing on estimated population-level impact of HIV prevention tools has been little studied, particularly in the context of HIV pre-exposure prophylaxis (PrEP).

PrEP with oral antiretrovirals is an effective prevention tool with potential for large population-level impact, especially when impact includes the indirect prevention benefits accrued by individuals not on PrEP.^2^ Most models of PrEP impact include heterogeneity in HIV-risk via heterogeneity in number of sexual partners,^3,4^ while some include assortative sexual mixing by attributes such as sexual activity level,^3^, age,^2–4^ and race/ethnicity.^2^

In the context of HIV epidemics among men who have sex with men (MSM), sexual mixing patterns also include seroadaptive behaviours like serosorting.^5^ Serosorting refers to preferential formation of partnerships between individuals of the same perceived HIV status.^5^ Data from behavioural surveys among MSM in high-income settings suggest that both HIV-positive and HIV-negative MSM practice serosorting as an HIV-prevention measure.^5,6^ However, across 15 transmission models of PrEP impact among MSM living in high-income settings (**Appendix 5-Table S5.1**), only three included serosorting.^2–4^ With the roll-out of PrEP across North America and Europe, data are emerging about potential changes in serosorting among MSM.^6,7^ PrEP may change serosorting patterns by reducing stigma and anxiety around sex with serodiscordant partners.^7^ Indeed, empirical data from a cross-sectional survey of MSM in Montréal, Canada demonstrate less population-level serosorting among HIV-negative MSM on PrEP compared with those who did not use PrEP.^6^

Mathematical models of PrEP impact among MSM have studied individual-level behaviour change among those on PrEP - often referred to as “risk compensation”. The models examined increases in number of partners,^3,8^ and reductions in condom use.^2–4,8^ Most models predicted that realistic increases in partner number or decreases in condom use would not fully offset, but could weaken, PrEP”s impact on reducing HIV transmission.^2–4,8^ To date, no models have explored the influence of serosorting on the estimated HIV transmission impact of PrEP at the population-level, or how PrEP impact could change if PrEP changes serosorting patterns.

We developed a mathematical model of HIV transmission among MSM living in Canadian urban settings. First, we compared the impact of PrEP under epidemics with serosorting to the impact of PrEP under comparable epidemics with sero-proportionate mixing. Second, under epidemics with serosorting, we compared the impact of PrEP under scenarios when PrEP-users stopped serosorting to scenarios when PrEP-users continued serosorting after starting PrEP.

## METHODS

### Model Overview

We developed a deterministic compartmental model of HIV transmission to reproduce the epidemiologic features of stable HIV epidemics among MSM living in the three largest Canadian cities (Montréal, Toronto, and Vancouver). The model includes five compartments defined by HIV status, HIV diagnosis, and the use of PrEP or antiretroviral treatment (ART) (**Figure 1, Appendix 1**). Individuals enter the model in the susceptible health-state at onset of sexual activity and exit the model due to death or cessation of sexual activity.

**Figure 1.**
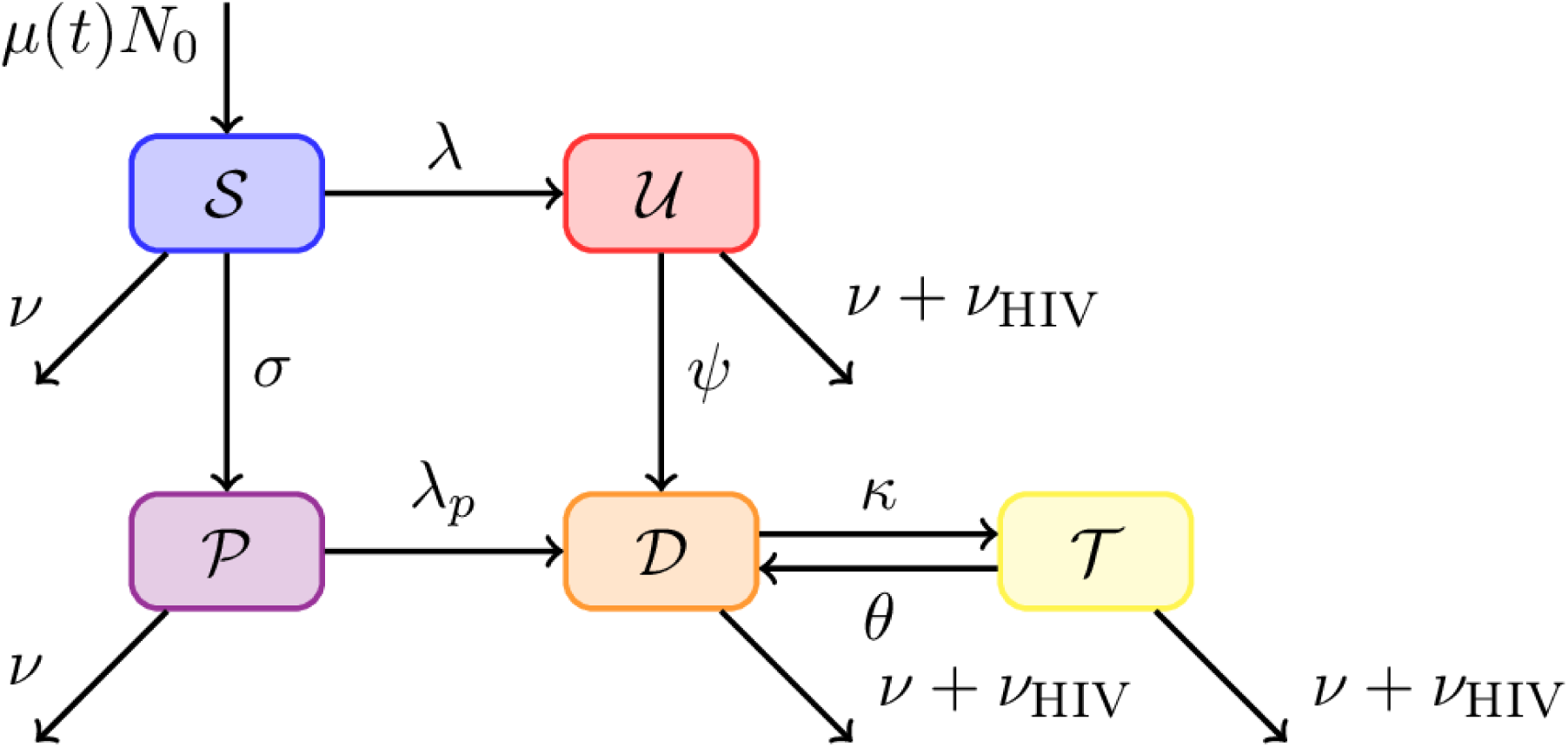
Schematic of the HIV transmission model structure with state transitions. Each box represents a health state with solid arrows for transitions between states, and exit due to cessation of sexual activity or mortality. Entry into the model incorporates population growth. S: susceptible, P: on pre-exposure prophylaxis, U: undiagnosed HIV, D: diagnosed HIV, T: on HIV antiretroviral treatment. A more comprehensive description of the model, with specific references to parameters used for each transition is provided in **Appendix 1.1**.

We sourced city/region or province-specific HIV surveillance reports and bio-behavioural surveys of MSM in Canada for estimates of HIV prevalence,^6,9–12^ annual new HIV diagnoses,^13–15^ and treatment parameters.^6,16,17^ We obtained demographic and sexual behavioural parameters from publicly-available behavioural surveys of MSM in Canada.^6,9,12^ We described the details of data sources and parameterization in **Appendix 2 and 3.2**.

We modelled HIV transmission via condomless receptive and insertive anal sex. The probability of HIV acquisition for a susceptible individual depended on per-act transmission probability of condomless insertive and receptive anal sex; number of sex partners; probability that the sex partner is living with HIV and not virally suppressed; number and type of anal sex acts per partnership; and condom use (**Table 1, Appendix 1.2**). We assumed 86% of MSM on ART achieved viral suppression (**Table 1**),^16^ those virally suppressed could not transmit HIV. Heterogeneity in HIV transmission risk was modelled via two sexual activity levels. The high activity group has 6 times as many sexual partners as the low activity group, to reproduce an incidence ratio of 6 between the high vs. low activity groups. The incidence ratio was informed by data suggesting that MSM with an HIV Incidence Risk Index score greater than or equal to 25 had six times the HIV incidence compared with those with a score less than 25 (**Appendix 2.2.2**);^18^ with the former comprising 6%-12% of the sexually active MSM population (prior range, detailed in **Appendix 2.2.3**).^19^ We applied the same rates of HIV testing and ART initiation in both groups, and proportionate mixing by sexual activity level.

**Table 1.**
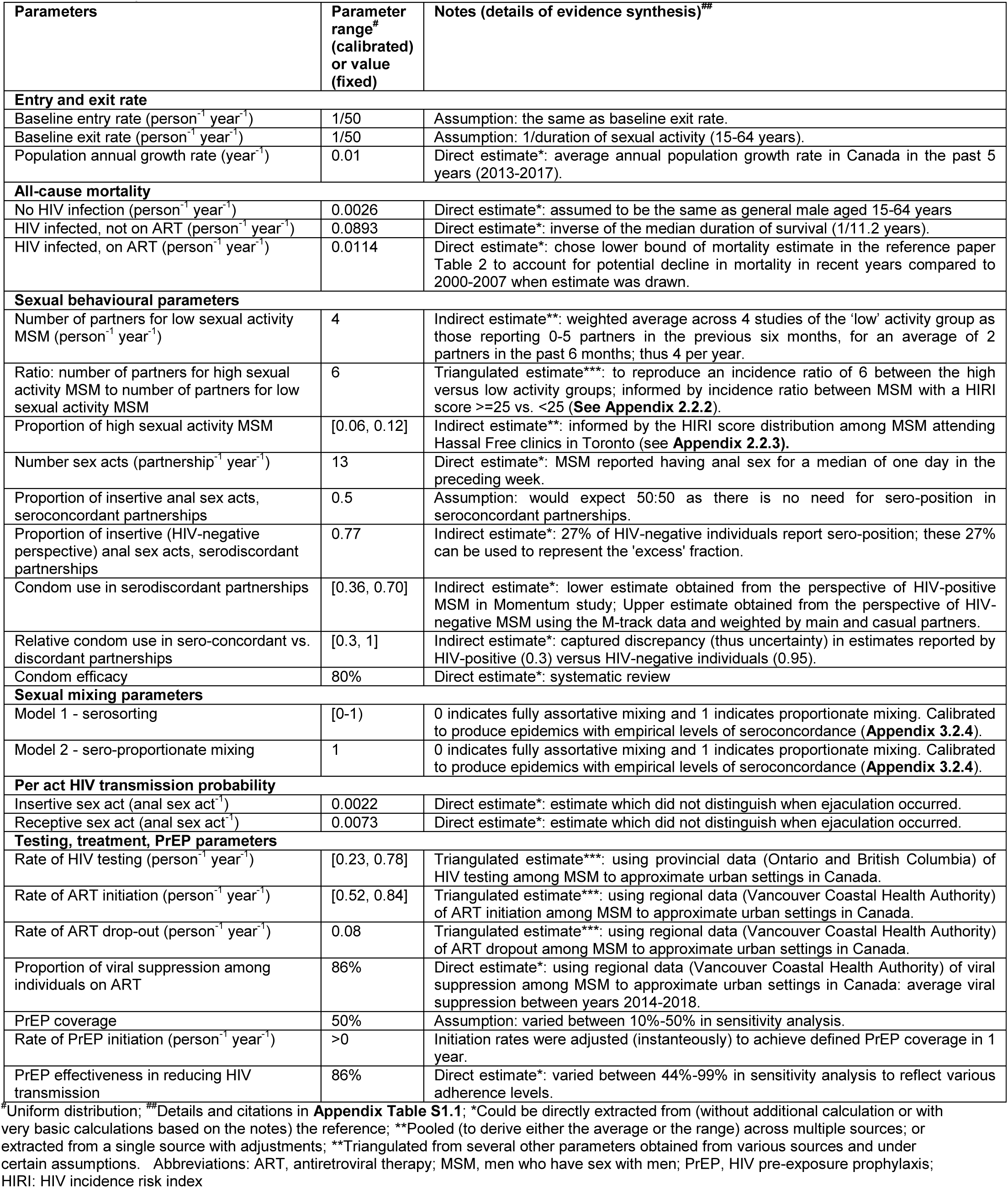
Model parameter values

We modelled sexual mixing by perceived HIV status via a parameter ϵ which controls the degree of assortative mixing, where value 0 indicates fully sero-assortative mixing, and value 1 indicates sero-proportionate mixing (selecting partners based on partnership availability by HIV status)(**Appendix 1.2.2**).^20^ We calibrated the value of ϵ within the range of 0 to 1 to fit to the empirical estimates of the population-level sexual mixing patterns by perceived HIV status (details below).

### Model calibration

We simulated and calibrated models separately under two assumptions: with serosorting vs. with sero-proportionate mixing. **Appendix 3** details the model calibration.

#### Model 1 - serosorting

We sampled 2000 sets of priors of the fitted parameters using Latin hypercube sampling,^21^ and calibrated the model to an equilibrium (**Table 1, Appendix 3.3.1**): HIV prevalence 10.3%-30.7%;^6,9–12^ annual number of new HIV diagnoses 94–909 per 100,000 MSM;^13–15^; and ART coverage among MSM diagnosed with HIV 81%-98%.^6,16,17^ We simultaneously calibrated our model to published, empirical estimates of two population-level seroconcordance values (**Appendix 3.2.4**): proportion of perceived seroconcordant partnerships by self-perceived HIV-negative individuals 83.3%-95.1%;^6,22^ and proportion of perceived seroconcordant partnerships by HIV-positive individuals 33.9%-76.5%.^6,23^ We retained 320 sets of calibrated posteriors for *Model 1*.

#### Model 2 - sero-proportionate mixing

For each set of fitted posteriors from the model under serosorting (*Model 1*), we obtained a matched set of posterior parameter values from the model with sero-proportionate mixing (*Model 2*) to generate comparable (<2% relative difference) HIV prevalence, new annual HIV diagnoses rate, and ART coverage. Thus, we set the value of ϵ =1 in *Model 2* reflecting sero-proportionate mixing. We then re-fit the two condom use parameters (condom use between perceived serodiscordant partnerships; and relative condom use in perceived seroconcordant vs. discordant partnerships) within their prior ranges in **Table 1**. For *Model 1* and *2* to generate the same HIV prevalence, something else must compensate for the difference in population-level HIV transmission risk changes in the absence vs. presence of serosorting. We selected condom use because of uncertainty surrounding its estimates, and because condom use can be considered a proxy for risk. We calibrated the two condom use parameters to fit *Model 2* to the matched equilibrium values of HIV prevalence, HIV new diagnoses rate and ART coverage generated by *Model 1* using an optimization algorithm (**Appendix 3.3.2**) and obtained 244 sets of calibrated posteriors for *Model 2*.

### PrEP intervention

#### Scenario 1 – PrEP did not modify sexual mixing patterns

After model calibration, we introduced PrEP intervention to both *Model 1* and *2*. We applied uniform access and uptake of PrEP by sexual activity level, with a linear increase in PrEP coverage until 30% coverage among HIV-negative individuals was achieved 1 year post-implementation. We varied coverage between 10%-50% in sensitivity analyses (**Appendix 2.6.2**). PrEP coverage remained stable thereafter, and we did not include PrEP discontinuation over the time-horizon of analyses for model simplification. We used PrEP effectiveness of 86% in our primary analysis, as per the IPERGAY study among MSM in France and Canada; and 44-99% effectiveness in sensitivity analyses.^24^

#### Scenario 2 – PrEP-mediated changes in serosorting

In *scenario 2*, we introduced changes in the serosorting patterns following PrEP initiation under the model with serosorting (*Model 1*), while maintaining all other elements of the PrEP intervention as with *scenario 1*. We assumed that 1) individuals on PrEP stopped serosorting (sero-proportionate partner selection) as soon as they initiated PrEP; 2) men not on PrEP adapted accordingly when they form partnerships with PrEP users to balance partnerships; and 3) men not on PrEP maintained the pre-intervention level of serosorting when they formed partnerships with other men not on PrEP. **Appendix 1.3.3** details the mathematical solutions to balancing partnerships while satisfying the above assumptions.

### Model Analyses

#### Influence of serosorting

We compared absolute difference in the relative HIV incidence reduction ten-year post PrEP intervention between *Model 1* with serosorting vs. *Model 2* with sero-proportionate mixing, under the PrEP intervention when PrEP did not change sexual mixing patterns (*Scenario 1*).

#### Influence of PrEP-mediated changes in serosorting

We used epidemics generated by *Model 1* to estimate the absolute difference in the ten-year relative HIV incidence reduction between two scenarios (counterfactuals): PrEP users stopped serosorting vs. continued serosorting.

#### Uncertainty and sensitivity analyses

To examine the influence of HIV epidemic features (HIV prevalence, fraction of undiagnosis, and ART coverage), and levels of serosorting on the results, we performed bivariate analyses using scatter plots and multivariable analyses using partial rank correlation coefficient (PRCC) to identify the most influential factors.^25^ We also examined a range of PrEP effectiveness (reflecting various adherence levels) and coverage levels to identify the intervention conditions under which serosorting and PrEP-mediated changes in serosorting would have the largest influence on PrEP impact.

### Role of the Funding Source

The funders of the study had no direct role in the conduct of the analysis, or in the study design, data collection, interpretation, or writing and submission of the report. The corresponding author had full access to the model and all the published data used in the modeling study and final responsibility for the decision to submit for publication.

### Ethics Statement

We only used published estimates to parameterize and calibrate the model (i.e. we did not access nor use individual-level datasets). Thus, the modeling study was exempt from institutional research ethics board review.

## RESULTS

### Model calibration

*Model 1* with serosorting reproduced the observed range of epidemics with respect to HIV prevalence (10.3%-24.8%), annual HIV diagnoses per 100,000 MSM (391-904), and ART coverage (82.5%-88.4%). By calibrating to empirical estimates on population-level seroconcordance measures from the perspectives of both HIV-negative and HIV-positive MSM, the posterior values of ϵ ranged from 0.29 to 0.81, reflecting various levels of serosorting. The estimated HIV incidence at equilibrium ranged from 0.5-1.7 per 100 person-years, HIV undiagnosed fraction ranged from 4.9%-15.8%, and all-cause mortality among individuals living with HIV ranged from 2.4-3.5 per 100 person-years. We present the kernel density-estimated distributions of all calibrated posteriors in **Appendix 4-Figure S4.1**.

*Model 2* with sero-proportionate mixing reproduced similar values of HIV prevalence, new HIV diagnosis rate, and ART coverage as *Model 1* with serosorting. To do so, the calibrated posteriors of condom use were higher in *Model 2* than in *Model 1* (**Figure 2**). Condom use had to be higher in *Model 2* because - given relatively low level of undiagnosed HIV (4.9%-15.8%) - epidemics with serosorting mean fewer partnerships where transmission could occur compared to epidemics with sero-proportionate mixing. For example, HIV-positive partners comprised 4.9%-16.7% of partnerships by HIV-negative individuals under serosorting vs. 14.1%-31.6% under sero-proportionate mixing (**Figure 2**). Thus, for *Models 1* and *2* to produce comparable epidemics, the per partnership transmission probability had to be higher in *Model 1 with serosorting* as reflected by lower condom use (**Figure 2**), compared to *Model 2* with sero-proportionate mixing.

**Figure 2.**
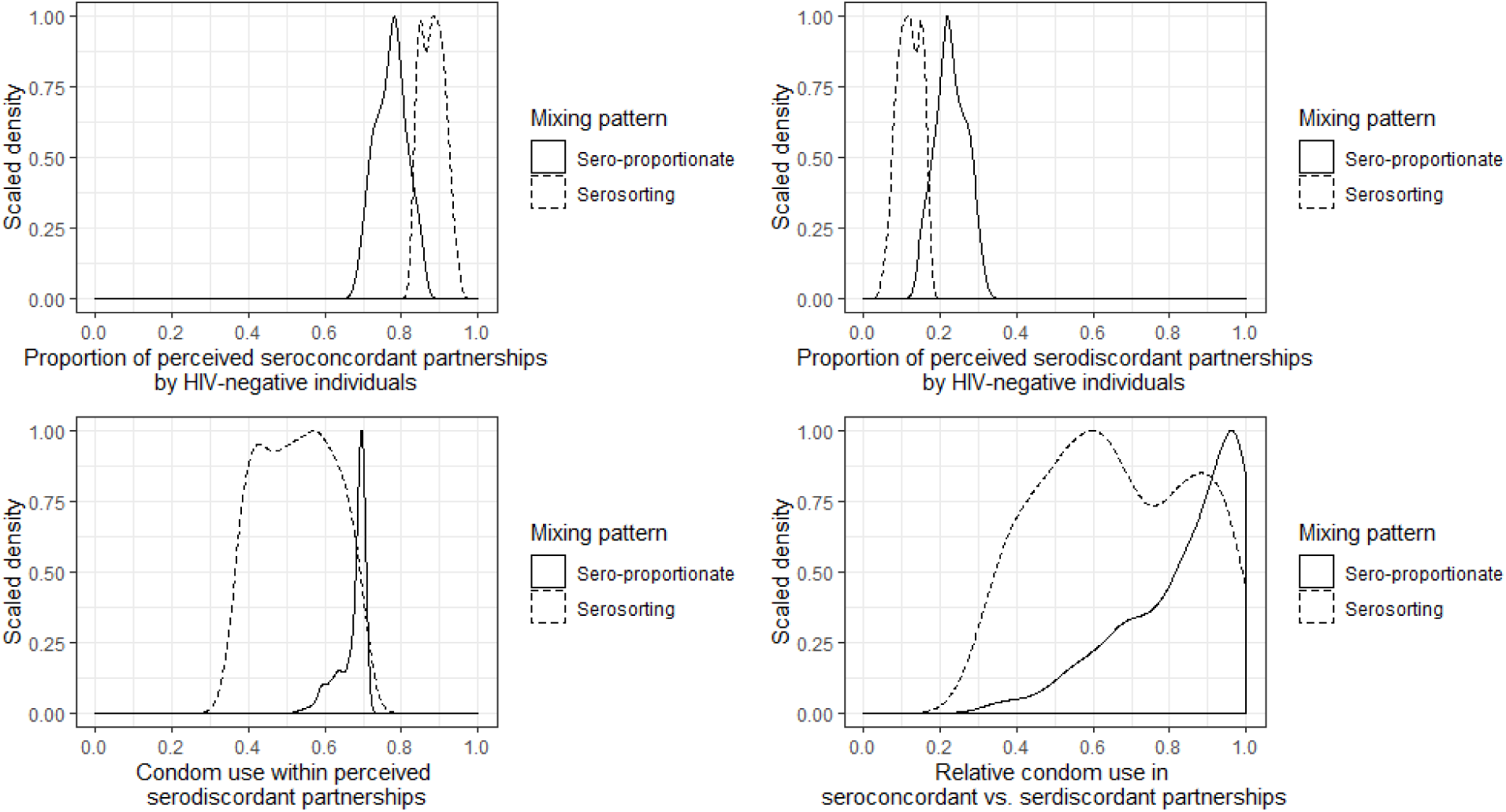
Compare the posterior partnership distribution by perceived HIV status, and posterior condom use in models with serosorting vs. models with sero-proportionate mixing.

### Influence of serosorting

*Model 1* with serosorting predicted a larger population-level HIV transmission impact of PrEP compared with *Model 2* with sero-proportionate mixing. As shown for one epidemic in **Figure 3(A)**, at 86% PrEP effectiveness and 30% coverage, the relative reduction in HIV incidence ten-year after PrEP initiation was 57.7% under serosorting and 44.7% under sero-proportionate mixing, reflecting an absolute difference of 13.0% in relative incidence reduction. Across all epidemics, the 10-year absolute difference in relative incidence reduction ranged from 2.0%-21.7% with a median of 9.5% (interquartile range: 6.7%-12.5%) when comparing serosorting to sero-proportionate mixing (**Figure 3(B)**). Variation in the magnitude of the influence of serosorting was largely explained by the level of serosorting, with a higher level of serosorting correlated with a larger influence (**Appendix 4-Figure S4.2**).

**Figure 3.**
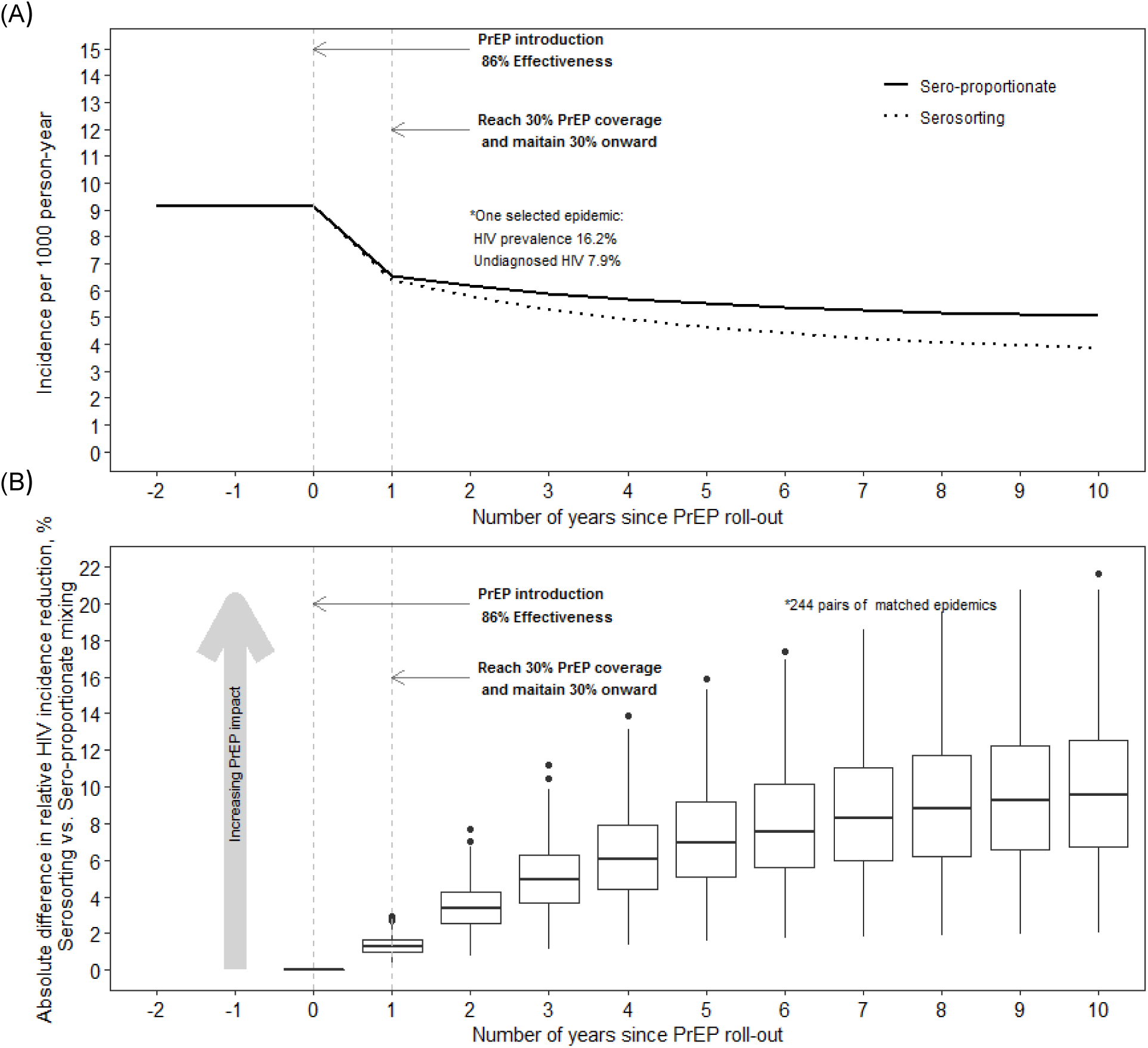
Comparison of pre-exposure prophylaxis (PrEP) impact on the population-level HIV transmission between models with serosorting vs. models with sero-proportionate mixing. (A) Incidence trajectory ten-year after PrEP introduction for one example epidemic. (B) Boxplots summary across 244 pairs of matched epidemics (match by HIV prevalence, new HIV diagnoses rate and antiretroviral treatment coverage) regarding the absolute difference in the relative HIV incidence reduction each year after PrEP introduction over ten-year period comparing serosorting to sero-proportionate mixing.

For a given PrEP coverage, the influence of serosorting on the transmission impact of PrEP decreased as PrEP effectiveness increased (shown using a single epidemic in **Figure 4**). This inverse relationship stems from a smaller marginal benefit at the individual-level from serosorting when individual-level PrEP effectiveness is high; and thus a smaller influence of serosorting at the population-level (**Appendix 4-Figure S4.3(A)**). The influence of serosorting was the largest at 50% coverage when PrEP effectiveness was low (44%); and peaked at 30% coverage when PrEP effectiveness was high (86%-99%)(**Figure 4**). This is because the rate of relative HIV incidence reduction due to PrEP diminishes when PrEP coverage exceeds 30%-50% (**Appendix 4-Figure S4.3(B)**).

**Figure 4.**
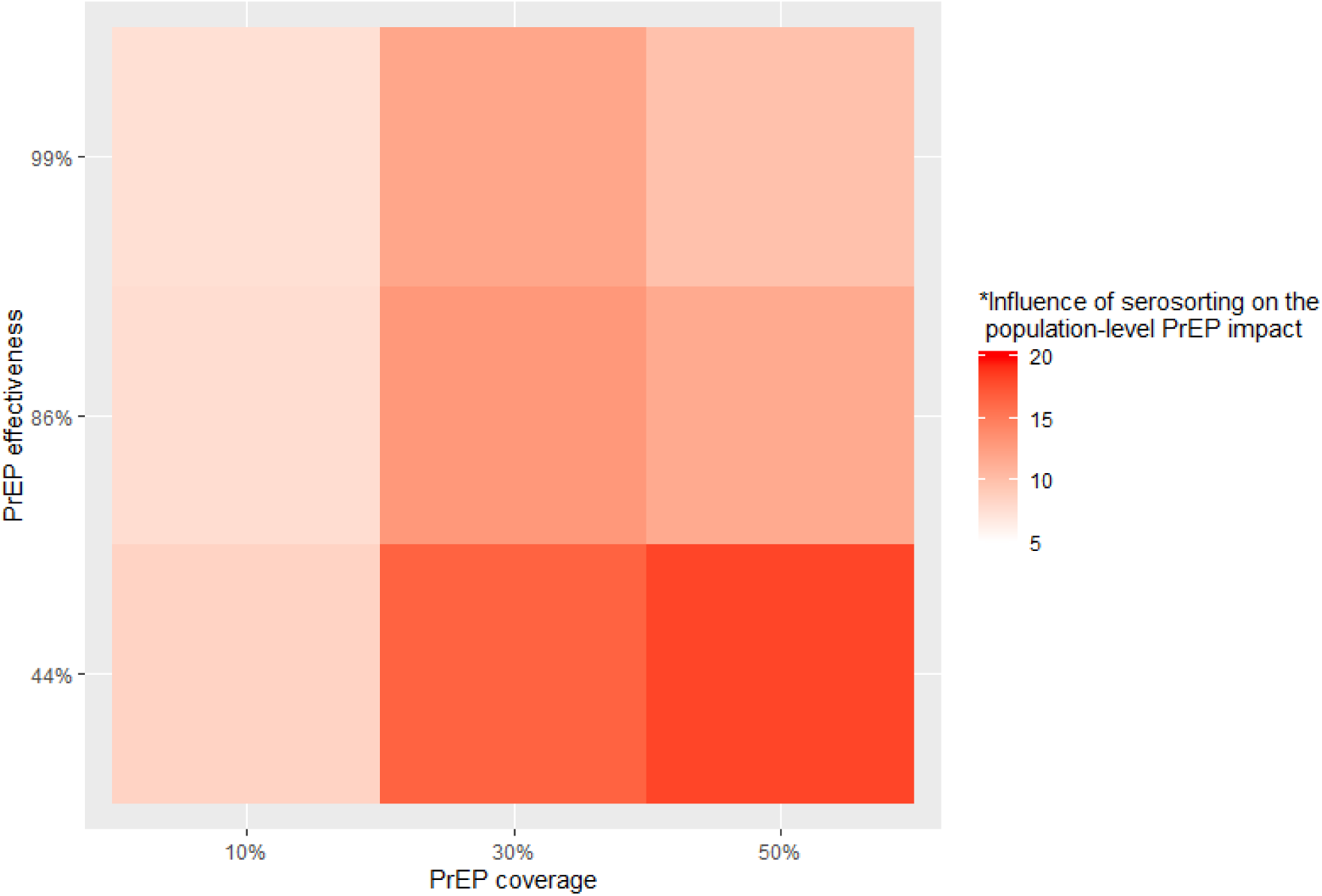
Variations in the influence of serosorting on the population-level HIV transmission impact of pre-exposure prophylaxis (PrEP), by PrEP coverage and effectiveness as demonstrated using one example epidemic (HIV prevalence 16.2%; undiagnosis fraction 7.9%). *Measured by absolute difference in the relative HIV incidence reduction ten-year after PrEP introduction between the model with serosorting vs. the model with sero-proportionate mixing.

### Influence of PrEP-mediated changes in serosorting

When PrEP users stopped serosorting there was a reduced impact of PrEP compared with scenarios when PrEP users continued serosorting (**Figure 5**). For example, at 86% PrEP effectiveness and 30% coverage, the absolute difference in relative HIV incidence reduction 10-year after PrEP initiation ranged from −7.2% to −1.1% between scenarios with and without PrEP-mediated changes in serosorting (**Figure 5**).

**Figure 5.**
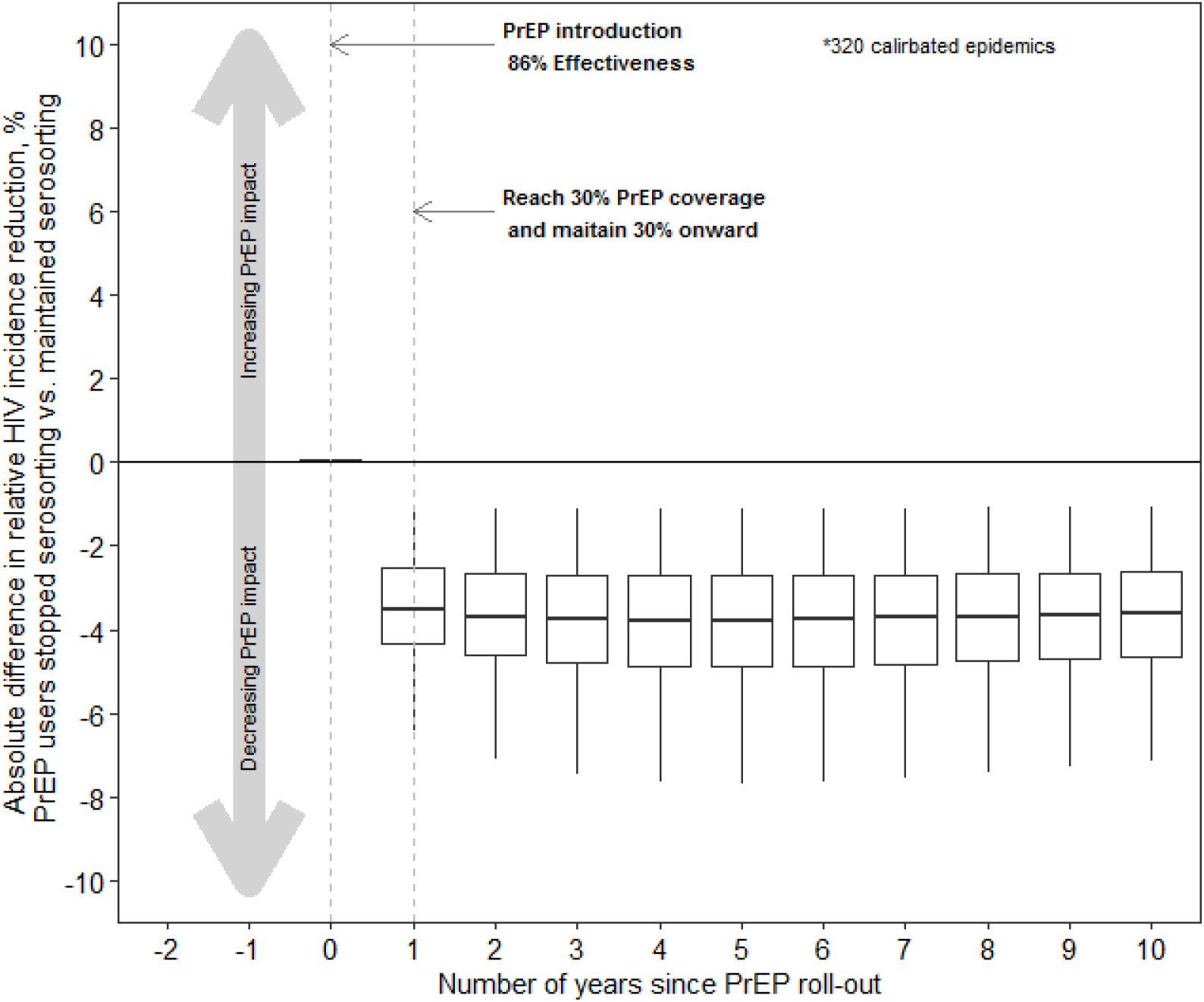
Comparison of pre-exposure prophylaxis (PrEP) impact on the population-level HIV transmission under scenarios when PrEP users stopped serosorting vs. scenarios when PrEP users continued serosorting. Boxplots summary across 320 epidemics regarding the absolute difference in the relative HIV incidence reduction each year after PrEP introduction over the ten-year period, between two scenarios.

In sensitivity analyses, the following factors demonstrated a strong association with the influence of PrEP-mediated changes in serosorting on PrEP impact (**Appendix 4-Table S4.1**): PrEP effectiveness (PRCC=0.91; positive association), level of serosorting (PRCC=-0.76; negative association), PrEP coverage (PRCC=-0.68, negative association), and pre-intervention HIV prevalence (PRCC=-0.37; negative association). As shown in **Figure 6**, when PrEP effectiveness was low (44%), PrEP-mediated changes in serosorting was more likely to reduce the transmission impact of PrEP, especially in settings with higher pre-intervention HIV prevalence, higher level of serosorting, and at higher PrEP coverage. However, when the effectiveness of PrEP was high (86%-99%), the influence of PrEP-mediated changes in serosorting had minimal influence on the transmission impact of PrEP (**Figure 6**).

**Figure 6.**
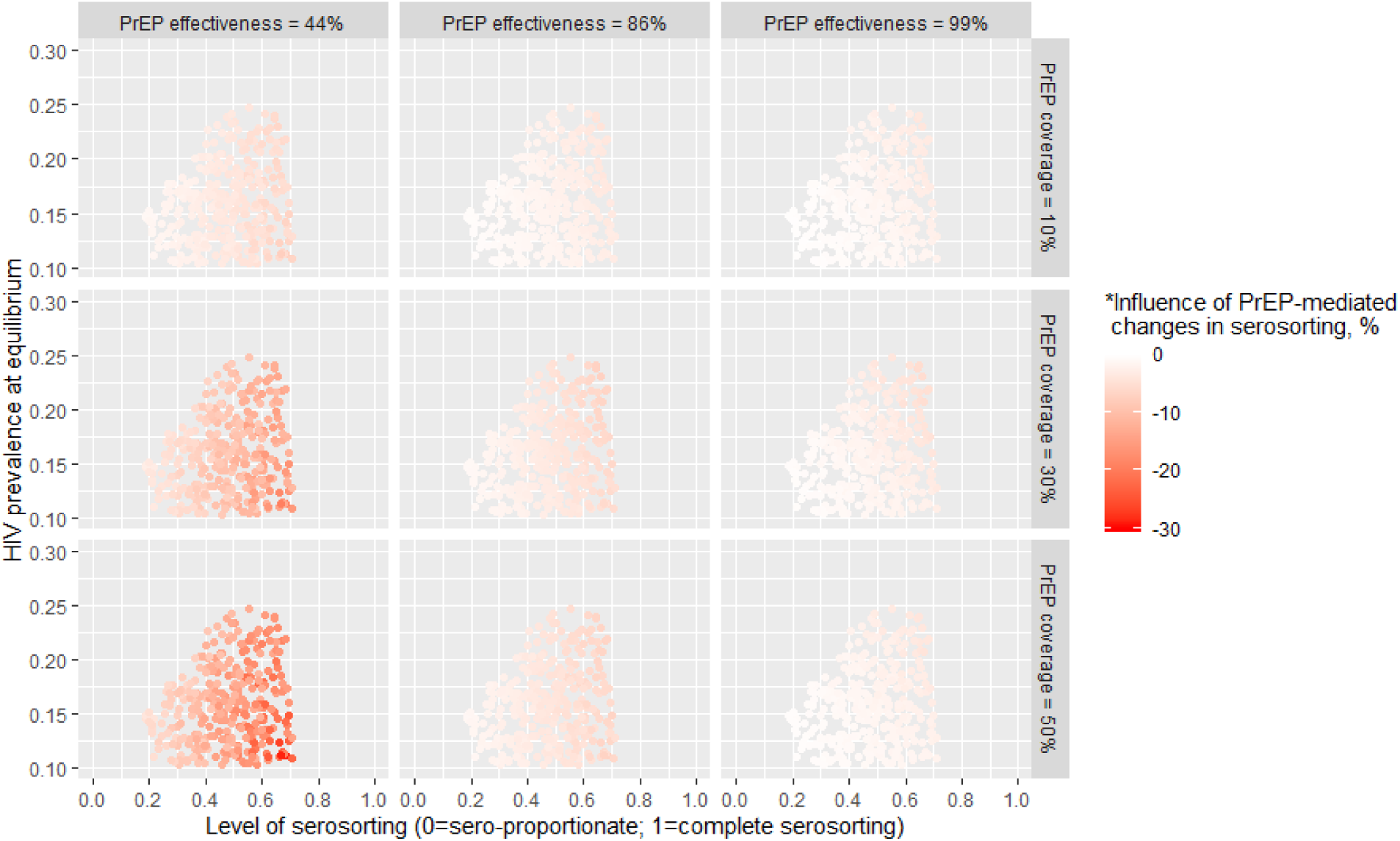
Variations in the influence of pre-exposure prophylaxis (PrEP)-mediated changes in serosorting on the population-level HIV transmission impact of PrEP by baseline level of serosorting, HIV prevalence at equilibrium, PrEP coverage, and effectiveness. *Measured by absolute difference in the relative HIV incidence reduction ten-year after PrEP introduction, comparing scenarios in which PrEP users stopped serosorting vs. maintained serosorting.

### Mechanism underlying PrEP-mediated changes in serosorting

Reductions in PrEP impact due to PrEP-mediated changes in serosorting can be explained by the nature of partnership balancing as shown in **Figure 7 (A)**, which compared the partnership distribution ten-year after PrEP initiation between scenarios when PrEP users stopped serosorting vs. continued serosorting. When PrEP users no longer serosort, their sexual partnerships comprise a higher proportion of HIV-positive partners, and thus a lower proportion of perceived HIV-negative partners (both on and not on PrEP)(**Figure 7 (A)**). Men not on PrEP (both perceived HIV-negative not on PrEP and HIV-positive) therefore also form partnerships with PrEP users in a sero-proportionate manner, in order to balance partnerships (mathematical proofs shown in **Appendix 1.3.3**). Consequently, the proportion of partnerships formed with PrEP users decreases for perceived HIV-negative individuals not on PrEP, and increases for HIV-positive individuals (**Figure 7 (A)**). Under the assumption that men not on PrEP continue to serosort when forming partnerships with other men not on PrEP, we provide the mathematical confirmation that the proportion of partnerships formed between HIV-positive and perceived HIV-negative individuals not on PrEP remained the same between both scenarios (**Figure 7 (A)**; proofs shown in **Appendix 1.3.3**). Finally, to satisfy partnership balancing overall, the proportion of perceived HIV-negative partners not on PrEP increases for perceived HIV-negative individuals not on PrEP, and the proportion of HIV-positive partners decreases for HIV-positive individuals (**Figure 7 (A)**).

**Figure 7.**
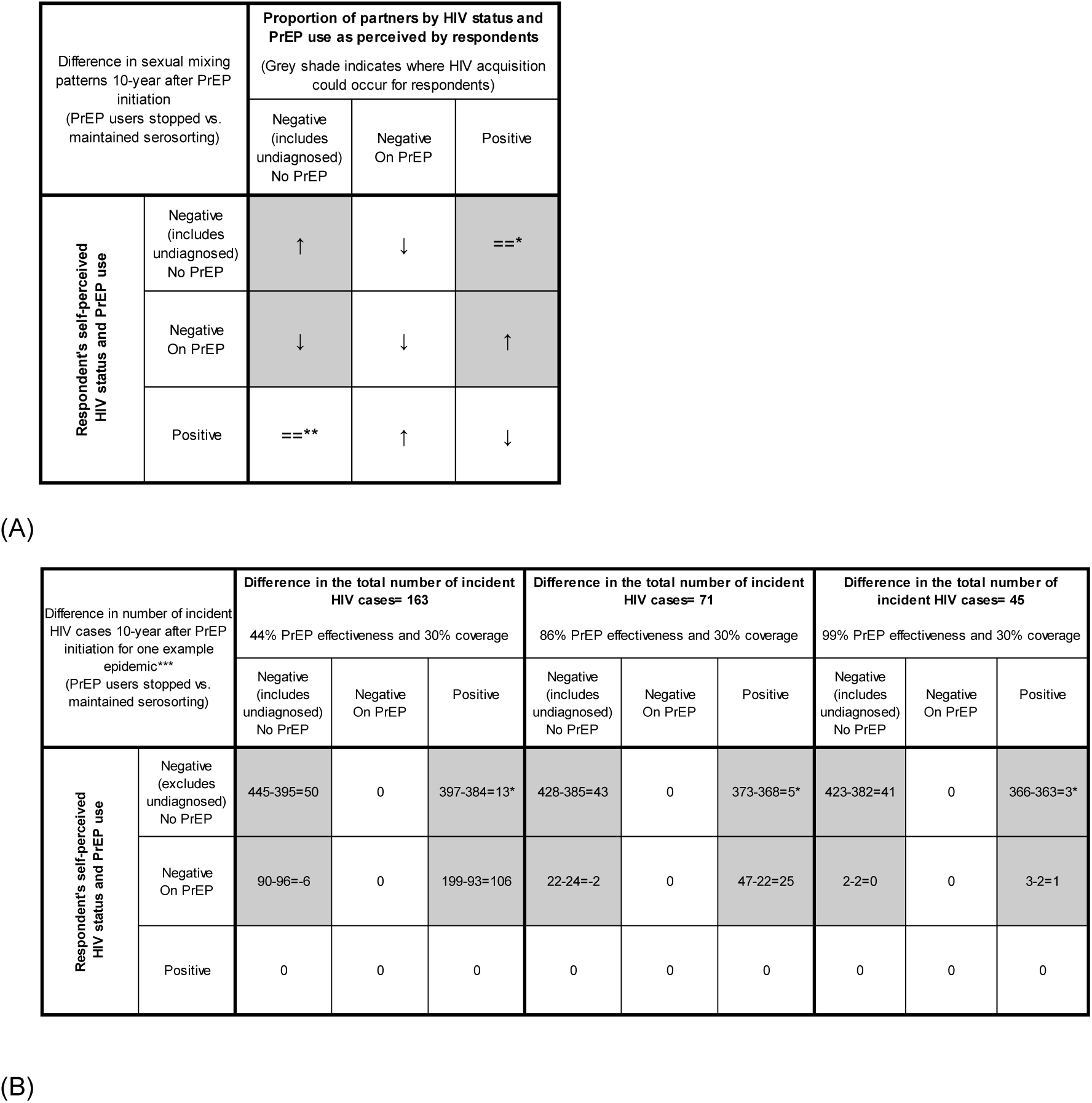
Demonstrating the influence of pre-exposure prophylaxis (PrEP)-mediated changes in serosorting (PrEP users stopped vs. maintained serosorting) on the population-level (A) sexual mixing patterns and (B) HIV transmission ten years after PrEP initiation. *Minor absolute difference of 0.02% to 0.56%; due to the fact that in scenarios when PrEP users stopped serosorting, the overall incidence reduction was smaller, and thus in the long term, there would be a slightly larger number of HIV-positive individuals, and a slightly smaller number of perceived HIV-negative individuals in the population, leading to a slightly higher proportion of HIV-positive partners, and a slightly lower proportion of perceived HIV-negative partners; **Minor absolute difference of −0.38% to −0.01%; details see notation*; ***Example epidemic reflects HIV prevalence 16.2%, and undiagnosis fraction 7.9%.

The difference in partnership distribution between two scenarios (PrEP users stopped vs. continued serosorting) meant that when we compared the number of incident infections ten-year into PrEP roll-out in the two scenarios, there were: fewer infections within partnerships between PrEP-users and their perceived HIV-negative (undiagnosed) partners (very small number of cases); more infections within partnerships between PrEP-users and their HIV-positive partners; and more infections within partnerships between HIV-negative individuals and their perceived HIV-negative (undiagnosed) partners (**Figure 7 (B)**). Therefore, there were more infections overall when PrEP users stopped serosorting vs. when PrEP users continued serosorting.

## DISCUSSION

Using a dynamic HIV transmission model among MSM, we found that patterns of sexual mixing influence the predicted population-level impact of PrEP on HIV incidence reduction. The impact of PrEP was higher in epidemics with serosorting, compared with comparable epidemics with sero-proportionate mixing. PrEP-mediated changes in serosorting (PrEP users stopped serosorting) reduced PrEP impact compared with scenarios when PrEP users continued serosorting; however reductions in PrEP impact were minimal if PrEP effectiveness was high. Only in the context of low PrEP effectiveness and high PrEP coverage do PrEP-mediated changes in serosorting have the potential to programmatically-meaningfully undermine the impact of PrEP.

Our findings suggest that in epidemic contexts where serosorting may reduce HIV transmission (i.e. settings with undiagnosed HIV <20% and ART coverage of >70%),^26^ models that ignore serosorting patterns (i.e. assume sero-proportionate mixing) could underestimate the projected transmission impact of PrEP, or overestimate the PrEP coverage required to achieve a desired population-level incidence reduction goal. Therefore, model-based evaluation of the impact of real-world PrEP implementation among MSM should incorporate serosorting patterns (which could also be seen as baseline risk reduction strategy employed by individuals), especially in high income-settings where the epidemics are similar to those examined in the current study. Indeed, our study also demonstrates that HIV transmission models can indeed be calibrated to patterns of serosorting measured at the population-level,^6^ to generate epidemics which reflect empirical levels of seroconcordance in sexual partnerships.

Our study is the first to our knowledge that directly examined the influence of PrEP-mediated changes in serosorting on the HIV transmission impact of PrEP and its underlying mechanism. To focus our hypothesis and examine the mechanisms by which differences in PrEP impact may be attributable to changes in patterns of sexual mixing mediated by PrEP, we purposefully designed our experiments to exclude other behavioural changes due to PrEP (e.g., reduction in condom use or increase in sexual partner numbers). We found that PrEP-mediated changes in serosorting had minimal overall influence on PrEP impact at the population-level when PrEP effectiveness was high. At all levels of PrEP effectiveness, there was a higher absolute number of incident HIV cases due to PrEP-mediated changes in serosorting for HIV-negative individuals not on PrEP via transmissions from partners living with undiagnosed HIV (**Figure 7(B)**). The modeled increase in infections was due to the downstream effects of PrEP-mediated changes in serosorting on the sexual network. Our findings highlight the importance of HIV testing to reduce the fraction or person-years of undiagnosed HIV in the population, especially after potential PrEP-mediated changes to sexual mixing.

PrEP-mediated changes in serosorting may considerably reduce PrEP impact if the PrEP effectiveness is low (44%) and as coverage reaches 30%. Thus, the influence of PrEP-mediated changes is relevant to the current state of PrEP roll-out in Canada, where by 2017-2019, PrEP coverage in Canadian cities is between 11%-23%.^27^ And although early data suggest high PrEP adherence (>95%), participants may be “early adopters” of PrEP whose high adherence may not represent the wider population of MSM.^28^ Indeed, in US cities with a longer history of PrEP roll-out suggest a high rate of PrEP cessation (62% of PrEP users discontinued PrEP over a median of 12-month follow-up);^29^ and if risk persists following PrEP cessation, then our lower bounds on 44% effectiveness is plausible when accounting for short-term adherence and long-term retention. Therefore, our findings support the need to monitor population-level sexual mixing patterns in addition to individual-level behavioural changes following PrEP initiation. Future studies should further examine the relationship between multiple behavioural changes (e.g., change in number of sex partners, condom use and mixing patterns) in PrEP users, and how they simultaneously influence the impact of PrEP.

Our study has several limitations. First, we restricted our study of PrEP-mediated changes in serosorting to a scenario where PrEP users stopped serosorting. Empirical data suggest less serosorting among PrEP users;^6^ thus our findings capture the maximum potential influence of PrEP-mediated changes in serosorting. Second, we simplified our PrEP intervention scenario (uniform access and uptake of PrEP by sexual activity level and stable PrEP coverage) and the influence of PrEP-mediated changes in serosorting are expected to vary with PrEP prioritized to the highest risk. Future analyses of PrEP-mediated changes in serosorting under different real-world PrEP intervention strategies is an important next step. Third, we did not include in our model additional HIV testing that could result from the implementation of PrEP program and thus potentially reducing undiagnosed HIV which in turn, could offset potential negative effects of PrEP-mediated changes in serosorting. Our rationale for its exclusion was based on empirical data suggesting few newly diagnosed HIV cases among individuals receiving HIV testing prior to PrEP initiation. Finally, as with many modelling studies, our findings are specific to the epidemiological context under study, and assuming PrEP interventions were initiated at an epidemic equilibrium.

In summary, transmission models that do not assess patterns of serosorting may underestimate the effectiveness of PrEP programs by underestimating the potential impacts of PrEP. Moreover, PrEP-mediated changes in serosorting could lead to programmatically-important reductions in PrEP impact which are similarly important to consider when planning and implementing PrEP. The findings highlight the importance of monitoring sexual mixing patterns and their changes over time in the design and evaluation of PrEP implementation.

## Contributors

SM, NM, and LW conceptualized and designed the study. LW and NM conducted evidence synthesis and parameterization. LW, AS, JK, and NM designed, modified, and analyzed the mathematical model. AS, JK, and LW conducted model coding, adaptation and calibration. LW, NM, AS, and JK designed and carried out the experiments. AS, HM, and SM contributed to evidence synthesis and parameter justification. LW, NM, and SM wrote the manuscript. LW, JK, NM, and AS wrote the appendix. All authors (LW, NM, AS, JK, HM, NJL, HLA, DHST, ANB, TAH, DMM, BDA, DRM, SB, and SM) provided critical input into decisions surrounding model structure, parameter justification, and the design of experiments. All authors (LW, NM, AS, JK, HM, NJL, HLA, DHST, ANB, TAH, DMM, BDA, DRM, SB, and SM) provided critical interpretation of results and critical manuscript review and editing.

## Declaration of Interests

There are no conflicts of interest to disclose.

## Data Availability

All model code (MATLAB) and parameter values can be found on our GitHub project repository. Data include: fixed parameter values, calibrated parameters: prior ranges and accepted posterior parameter sets, and calibration targets: acceptable ranges and accepted posterior predicted values. Values of model parameters (inputs) and calibration targets (outputs) were estimated from the data sources described in Table S2.1.

https://github.com/mishra-lab/prep-serosort

## Acknowledgments

This study was funded by the Canadian Institutes of Health Research (CIHR) foundation grant FN-13455.

SM and DHST are supported by a CIHR and the Ontario HIV Treatment Network (OHTN) New Investigator Award. TAH is supported by an OHTN Applied HIV Research Chair Award. DMM and NJL are supported by Scholar Awards from the Michael Smith Foundation for Health Research (#5209, #16863). NM was supported by the CIHR-funded Canadian HIV Trials Network Postdoctoral Fellowship. We would like to thank Kristy Yiu for supporting submission and project coordination, and Steven Tingley for helpful discussions surrounding model structure.

Some of the model parameters in the current modeling paper drew on estimates published in Wang et al. 2019 (https://doi.org/10.1093/aje/kwz231).We acknowledge the Engage study and its funders (Canadian Institutes of Health Research (CIHR) Team Grant [TE2-138299]; CIHR Canadian HIV Trials Network [CTN 300]; Canadian Foundation for AIDS Research [Engage]; Canadian Blood Services [MSM2017LP-OD]; Ontario HIV Treatment Network (OHTN) [1051]; Ryerson University [no related grant number]; and Public Health Agency of Canada [4500370314]) which supported the independently published results in Wang et al. 2019 (https://doi.org/10.1093/aje/kwz231).

## Notes

### Competing Interest Statement

The authors have declared no competing interest.

